# Evaluating efficiency of pooling specimens for PCR-based detection of COVID-19

**DOI:** 10.1101/2020.05.02.20087221

**Authors:** Supaporn Wacharapluesadee, Thongchai Kaewpom, Weenassarin Ampoot, Siriporn Ghai, Worrawat Khamhang, Kanthita Worachotsueptrakun, Phanni Wanthong, Chatchai Nopvichai, Thirawat Supharatpariyakorn, Opass Putcharoen, Leilani Paitoonpong, Gompol Suwanpimolkul, Watsamon Jantarabenjakul, Pasin Hemachudha, Artit Krichphiphat, Rome Buathong, Tanarak Plipat, Thiravat Hemachudha

**Author notes:** **CORRESPONDING AUTHOR:** Name: Supaporn Wacharapluesadee, Address: Thai Red Cross Emerging Infectious Diseases Health Science Centre, World Health Organization Collaborating Centre for Research and Training on Viral Zoonoses, King Chulalongkorn Memorial Hospital, Faculty of Medicine, Chulalongkorn University, Rama IV Road, Pathumwan, Bangkok 10330 Thailand.

## Abstract

In the age of a pandemic, such as the ongoing one caused by SARS-CoV-2, the world faces limited supply of tests, PPE and reagents, and factories are struggling to meet the growing demands. This study aimed to evaluate the efficacy of pooling specimen for testing of SARS-CoV-2 virus, to determine whether costs and resource savings could be achieved without impacting the sensitivity of the testing. Ten specimens were pooled for testing, containing either one or two known positive specimen of varying viral concentrations. Pooling specimens did not affect the sensitivity of detecting SARS-CoV-2, and the PCR cycle threshold (Ct) between testing of pooling specimen and subsequent individual testing was not significantly different using paired t-test. This study also identified cost savings garnered from pooling of specimen for testing at 4 differing prevalence rates, ranging from 0.1-10%. Pooling specimens to test for COVID-19 infection in low prevalence areas or in low risk population can dramatically decrease the resources burden on lab operations by up to 80%. This paves the possibility for large-scale population screening, allowing for assured policy decisions by governmental bodies to ease lockdown restrictions in areas with low incidence of infection, or with lower risk populations.

## INTRODUCTION

The ongoing COVID-19 pandemic has highlighted the need for early diagnosis of emerging infectious diseases to better contain an outbreak. Testing for SARS-CoV-2, the virus that causes COVID-19, has been limited due to factors including high cost and low availability of reagents, lack of personal protective equipment (PPE) and other consumables, and the sheer volume of samples be tested. Factories have been struggling to meet the growing demands for these necessities^1,2^. To date, countries that are able to screen patients swiftly have fared better in containing the COVID-19 outbreak and suppressing the mortality rate associated with the disease^3^. The rapid diagnosis of COVID-19 in both symptomatic and asymptomatic patients can shed light on transmission patterns and facilitate contact tracing^2,3^. Large scale population screening for COVID-19 infection is generally considered a necessary part of an exit strategy from the coronavirus lockdown, and in reinforcing infection control measures in hospitals and among health care workers.

Specimen pooling is a method of screening large number of patients for an infection, and typically involves combining multiple patient specimens into a single test sample, then testing multiple such samples. This approach has the advantage of cost-effectiveness and speed, and was used to retrospectively screen for COVID-19 in specimens that were negative for common respiratory viruses earlier in the course of the pandemic in the United States^4^, and in France to rapidly screen multiple returning expatriates from China with low suspicion of infection^5^. Specimen pooling has also been used in screening efforts for several other infectious diseases, including donated blood samples for HIV^6–9^.

Pooling nasopharyngeal and throat swab (NT) specimens would be more economical than individually testing all specimens from low-risk populations, particularly in limited-resource settings^10^. The current study was undertaken to compare laboratory results from sample pooling (10 samples) with the standard real-time polymerase chain reaction (qPCR) testing without pooling, as a means of preparing for a broad pooling-based screening effort to ensure that detection accuracy will not be compromised.

## MATERIALS AND METHODS

This study is an evaluation of laboratory techniques using archived clinical specimens and was exempted from Chulalongkorn University Institutional Review Board (IRB) review. NT specimens used in this study had been collected from patients under investigation (PUI) for COVID-19 infection at King Chulalongkorn Memorial Hospital, placed in viral transport media (VTM) and sent to the Thai Red Cross Emerging Infectious Diseases Health Science Centre Laboratory for testing between February 1, and March 31, 2020. All specimens had been stored at −80°C. In addition, 50 SARS-CoV-2 negative NT specimens in VTM from routine diagnoses (1.0 mL each), as determined by real-time PCR (BGI, Shenzhen, China), were pooled, and this pooled negative NT-VTM served as the negative portion of all samples tested (Figure 1). These negative pooled specimens were re-tested for SARS-CoV-2 using real-time PCR to confirm the negative result prior to pooling with selected positive specimens.

**Figure 1.**
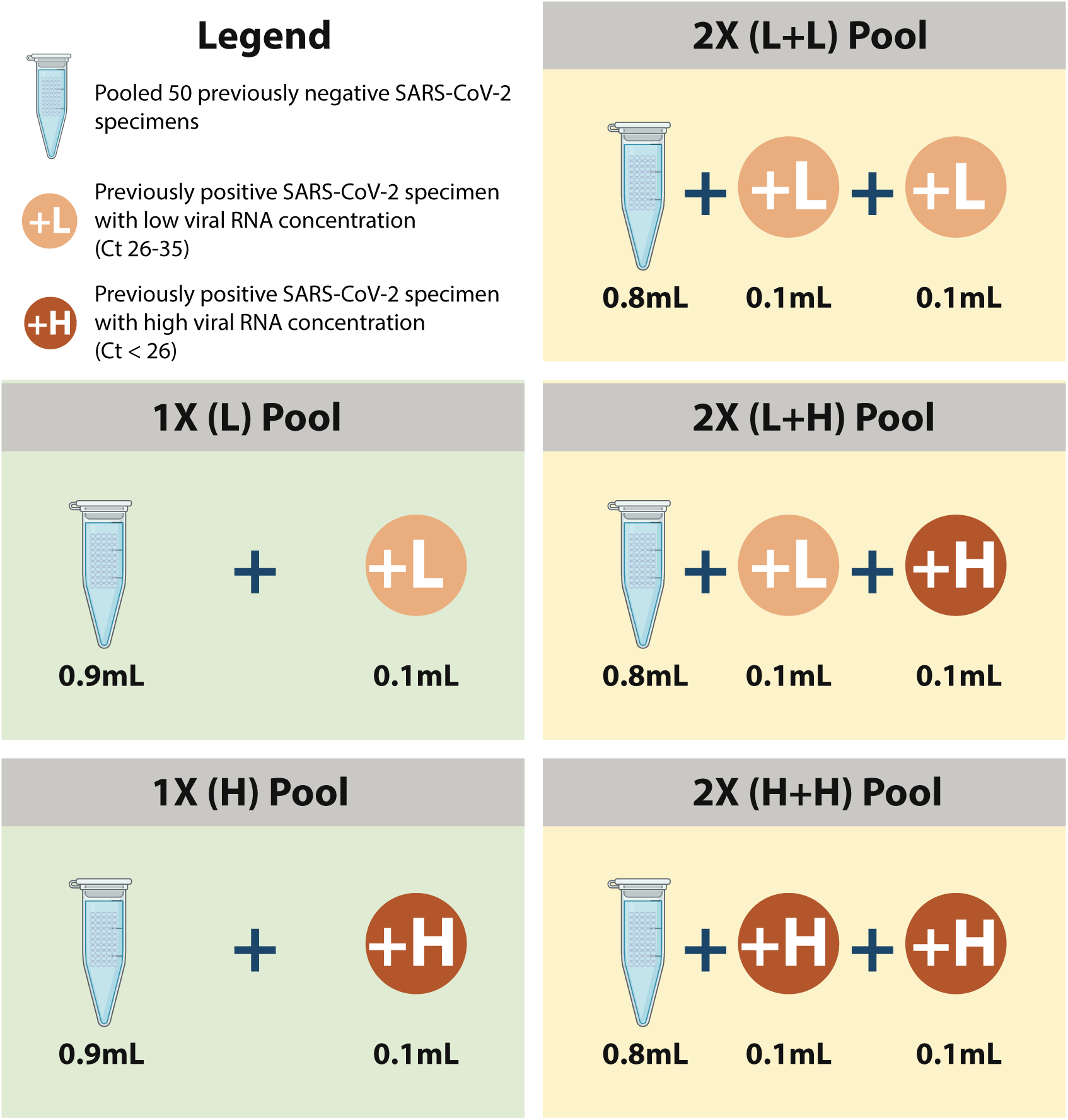
Illustrates the experimental design of the pooling strategies tested in this study

This study used magnetic extraction-based assay (NUCLISENS^®^, easyMag^®^, bioMérieux, Marcy-l’Étoile, France) based on the Boom method to extract DNA and RNA, which allows a maximum specimen volume of 1.0mL^11^. By using magnetic beads to capture DNA and RNA during the extraction step, pooling 10 specimens of 0.1mL each (total of 1.0 mL extraction sample) can result in the same extraction capability as 0.1mL if the elution volume at the end is equal and there is no PCR interference from the specimen such as lipid, protein or cell debris.

Two pooling ratios were evaluated in this study, termed 1X and 2X. In the 1X ratio, 0.1mL of NT-VTM from one SARS-CoV-2 positive specimen was combined with 0.9 mL pooled negative NT-VTM, thus modeling a 10% infection rate. Correspondingly, in the 2X ratio, 0.1mL of NT-VTM each from two SARS-CoV-2 positive specimens were pooled with 0.8 mL pooled negative NT-VTM, thus modeling a 20% infection rate (see Figure 1). All 1.0 mL pooled samples in this study were then processed for nucleic acid extraction using NUCLISENS^®^ easyMAG^®^ instrument (bioMérieux). In addition, 0.1mL of the same positive specimens that were used in the pooled samples were re-tested for sensitivity comparison using a separate extraction system (EZ1, Qiagen, Hilden, Germany). Real-time PCR (qPCR) for detection of SARS-CoV-2 was performed using a commercial kit which targets the ORF1ab gene as per the manufacturer’s protocol (BGI, Shenzhen, China). The protocol’s stated limit of detection of ORF1ab real-time PCR was 100 copies/mL and the cutoff PCR cycle threshold (Ct) was 38.

Previously positive specimens with high and low-concentrations of RNA, as determined by PCR cycle threshold (Ct) values at the time of detection, were selected to determine the effect of viral load on pooling to ensure sensitivity and accuracy of the assays is maintained (Table 1). Low Ct values indicate the presence of higher amounts of viral RNA and high Ct values indicate lower amounts. In this study, specimens with Ct values between 26 – 35 were considered to have low concentrations of viral RNA, while those with Ct values lower than 26 were considered to have of high-concentrations viral RNA. The experimental design is summarized in Figure 1.

**Table 1.**
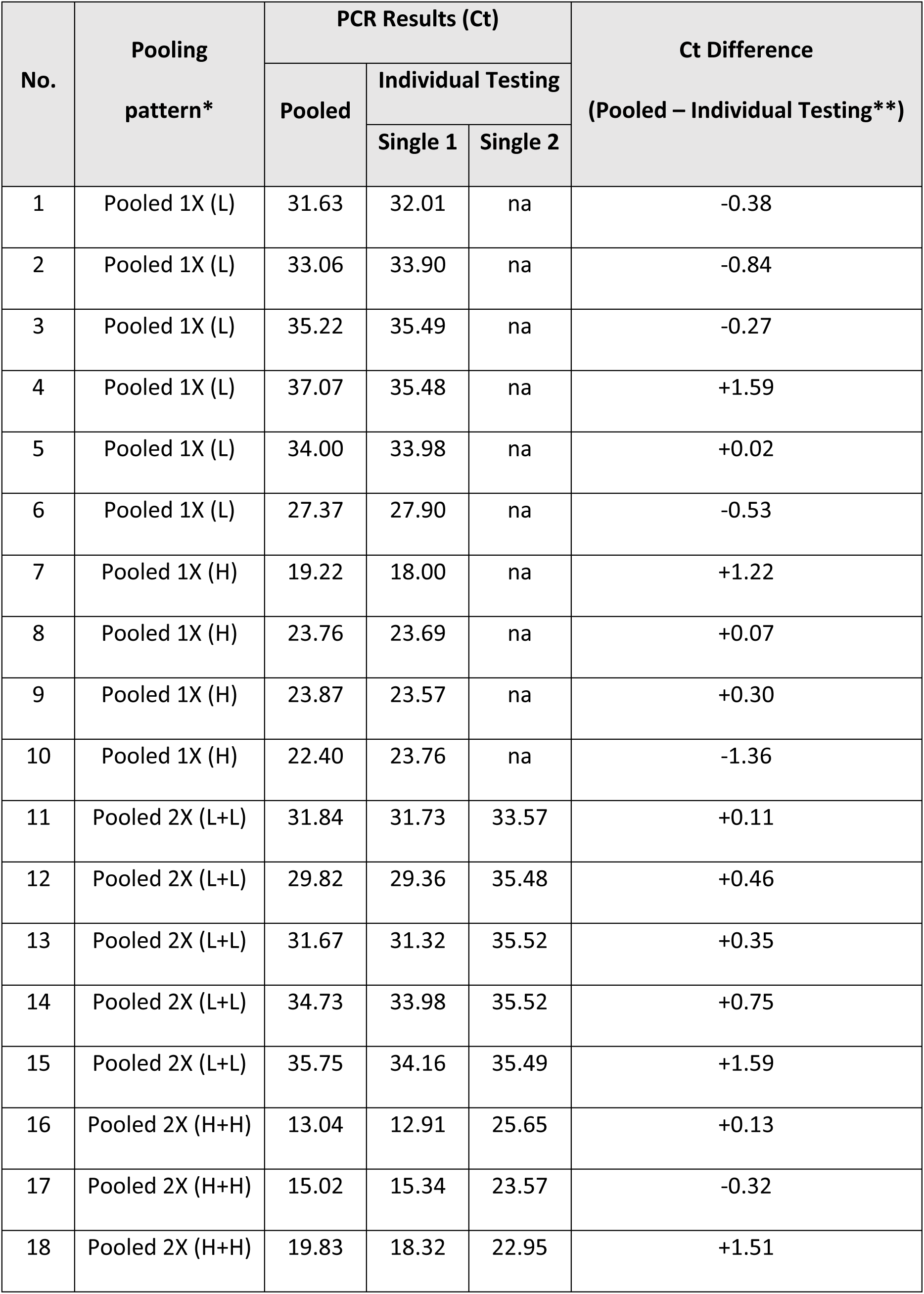
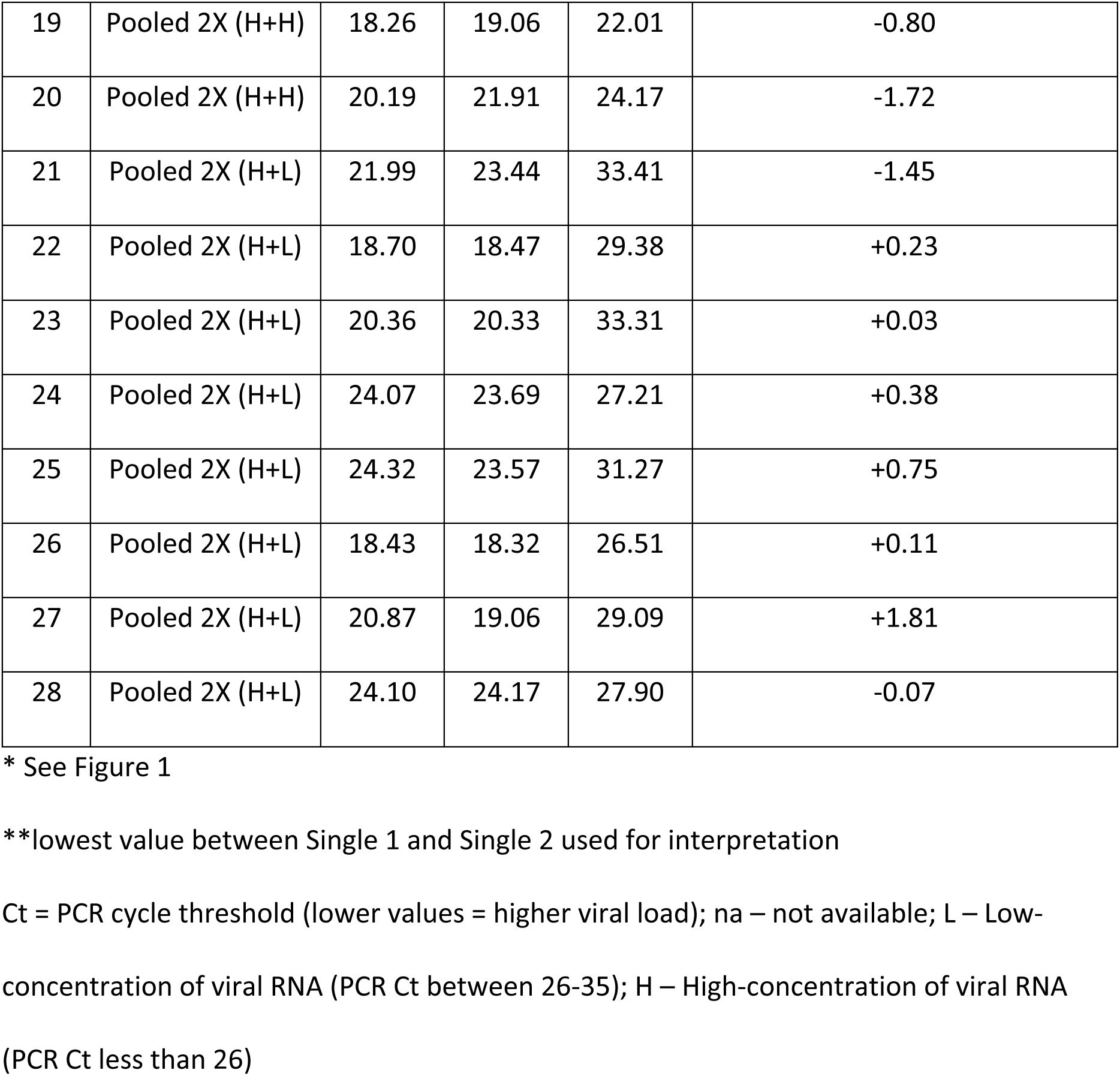
The PCR results of 1x (one positive in 10) and 2x (two positive in 10) pooled specimens and individual NT specimens are demonstrated as PCR Ct value

Twenty-eight 10-specimen pools were prepared. Ten of these had a 1X pooling ratio and 18 had 2X ratios. Among the 1X ratio, six had low viral concentrations, (L, Ct values from 27.90 to 35.49) and four had high viral concentrations (H, Ct values from 18.00 to 23.76). The 2X ratio pools had two positive specimens each, with viral concentrations as follows: five pools had two low-concentration specimens (L+L, Ct values from 29.82 to 35.52), five pools had two high concentration specimens (H+H Ct values from 12.91 to 25.56), and eight pools had one high and one low concentration specimens (H+L, Ct values from 18.47 to 33.41).

The sensitivity of viral RNA detection for each pool was compared with the sensitivity of PCR results for the individually tested positive specimen in that pool. For 2X ratio pools, the positive specimen with the lower Ct value, when individually tested, was used for comparison (Table 1).

## RESULTS

All 1X ratio pools were positive, with Ct value difference within a range of −1.36 to +1.59 when compared to individual (non-pooled) testing. Negative and positive values of Ct indicates higher and lower sensitivity of pooling, respectively. All 2X ratio pools were positive, with Ct value difference within a range of −1.72 to +1.81 when compared to individual (non-pooled) testing (Table 1). Statistical paired t-test was calculated to compare the Ct value differences between pooled (including all patterns in Figure 1) and individual tests. The result showed no significant difference in all comparisons including individual vs 1X L ratio pool (p = 0.853), or individual vs 1X H ratio pool (p = 0.921). The 2X pooling ratio showed similar results. There were no significant difference between the Ct values of individual testing vs ratio pools 2X L+L, 2X H+L, or 2x H+H (p = 0.063, 0.507, and 0.6766, respectively). Thus, sensitivity was not affected by pooling specimens, regardless of viral load, while accuracy was maintained.

Cost effectiveness of the pooling strategy was calculated, based on varying disease prevalence rates (0.1-10%) (Table 2). Pooling appears most cost-effective when testing among populations with lower COVID-19 prevalence. Estimated laboratory costs were reduced from $35 per patient to $3.85, $6.85, $17.54, $26.30 at prevalences of COVID-19 in the tested population of 0.1%, 1, 5%, and 10% respectively. By this estimation, pooled-specimen testing of 1,000,000 subjects in a population with 1% COVID-19 prevalence would save approximately $28.15 million, assuming evenly distributed positive specimens in each pool (Table 2).

**Table 2.** Cost comparison for pooled qPCR for 4 different prevalence rates

| Total population | 1,000,000 samples |  |  |  |
| --- | --- | --- | --- | --- |
| % infection | 0.10% | 1.00% | 5.00% | 10.00% |
| % of the non-infected samples | 99.90% | 99.00% | 95.00% | 90.00% |
| Number of samples per pool | 10 | 10 | 10 | 10 |
| Total number of pool | 100,000 | 100,000 | 100,000 | 100,000 |
| % of pool with no infection* | 99.00% | 90.44% | 59.87% | 34.87% |
| Total number of pool without an infection | 99,004 | 90,438 | 59,874 | 34,868 |
| Total number of pool with an infection | 996 | 9,562 | 40,126 | 65,132 |
| Number of samples that need to be tested individually after pooled PCR | 9,955 | 95,618 | 401,263 | 651,322 |
| Total number of tests that need to be performed | 109,955 | 195,618 | 501,263 | 751,322 |
| Cost per test (USD) | \$35.00 | \$35.00 | \$35.00 | \$35.00 |
| Total cost of individual testing | \$35,000,000.00 | \$35,000,000.00 | \$35,000,000.00 | \$35,000,000.00 |
| Total cost of pooling specimens | \$3,848,429.19 | \$6,846,627.37 | \$17,544,207.13 | \$26,296,254.60 |
| % discount | 89.00% | 80.44% | 49.87% | 24.87% |
| Cost per patient | \$3.85 | \$6.85 | \$17.54 | \$26.30 |

\*% of pool with no infection = (% of non- infected samples in one pool)^number of samples
per pools

## DISCUSSION

This study demonstrates that pooling specimens does not compromise the sensitivity of detecting SARS-CoV-2, regardless of viral load. The lowest viral concentration used in this study was at Ct 35.49 which was detected from both pooled and individual testing. Ct value more than 35 was considered weakly positive, and the effect of pooling on these samples will need to be further studied. In 2X ratio pooling, RT PCR testing detected higher viral concentrations (lower PCR Ct) compared to those of the corresponding positive specimens when tested individually. This suggests that pooling specimens did not lower the sensitivity of PCR testing but actually increased the viral concentration when more than one positive sample was present in the same pool which combined the viral amount from 2 samples in the same extraction tube. The nucleic extraction system used in this study allowed nucleic acid extraction from 1.0 mL of NT specimens without reducing the sensitivity as compared to 0.1 mL of individual extraction. Further, similar PCR Ct values (within +/− 2 Ct; statistically not significant) between pooled and non-pooled specimens indicated there was no interference of PCR inhibitor from 1.0 mL pooled specimens in one extraction tube.

Beyond maintaining accuracy, specimen pooling will almost certainly reduce cost. For example, if 1% of the population is infected, pooling 10 specimens can reduce the cost of laboratory operation by about 80% (Table 2). However, in the case of 10% prevalence, pooling specimens will only save 24.87%, as positive pooled samples will need to be individually tested. Thus, this is especially useful in areas with low prevalence rates, or when conducting pro-active surveillance in areas of low infection rate. Proactive surveillance, particularly in asymptomatic cases, remains a challenge to overcome in order to exit lockdown, where screening on a massive scale is required.

A previous study found that pooling at ratio of 1 to 5 (50 μL of each specimen for total of 250 μL pooled extraction) retained accuracy of the test and resulted in greater efficiency of test resources^10^ as well as demonstrating that when the prevalence of COVID-19 is 1%, the optimal specimen pool size is 11 with an overall increase in testing efficiency calculated at 400%. In this study, a 10-specimen pool size (100 μL each specimen) was chosen based on the capacity of the RNA extraction system in the laboratory where this study was performed, and the result was similar to 5 samples pooling^10^.

A limitation of this study is the maximum number of two positive specimens in the 10-specimen pool. In theory, more positive specimens in a pool could decrease the sensitivity of qPCR as it would result in too many viral copies, causing an insufficiency of PCR enzyme and other reagents in the mix to amplify all the viral copies. Practically, however, this does not affect the overall testing results, since positive pools would require individual testing in any case.

Rapid identification of SARS-CoV-2 infection is crucial to curb the COVID-19 pandemic. The current gold standard for testing SARS-CoV-2 is real-time PCR, which requires resources that are currently limited, along with specialized equipment and technically skilled labor. Shortage of testing reagents and equipment in countries where there is no capability to produce their own tests may result in delays in testing and result in reduced effectiveness in containing the outbreak. Pooled specimen testing would enable substantial savings in reagent costs, technical burden and time to generate laboratory results.

## Data Availability

All data referred to in this manuscript is included in the manuscript.

## ACKNOWLEDGEMENTS

This study was supported by a research grant from the King Chulalongkorn Memorial Hospital’s Excellent Center Program, National Research Council of Thailand (NRCT), and the Biological Threat Reduction Program (BTRP) of the US Defense Threat Reduction Agency (DTRA). We would also like to acknowledge Dr. Paul Gaudio (Yale University) for his kind assistance in the critical editing of this manuscript.

